# Quantification of specific antibodies against SARS-CoV-2 in breast milk of lactating women vaccinated with an mRNA vaccine

**DOI:** 10.1101/2021.04.05.21254819

**Authors:** Erika Esteve-Palau, Araceli Gonzalez-Cuevas, M. Eugenia Guerrero, Clara Garcia-Terol, M. Carmen Alvarez, Geneva Garcia-Aranda, David Casadevall, Vicens Diaz-Brito

## Abstract

The recent approval of vaccines against COVID-19 has generated great concern among breastfeeding women, since these patients were excluded from vaccination clinical trials. The present study aimed to analyze the levels of specific SARS-CoV-2 antibodies in breast milk of mRNA-vaccinated women across time and their correlation with serum antibody levels.

**Methods:** prospective study including lactating women aged over 18 who were vaccinated against SARS-CoV-2 with the Pfizer-BioNTech® COVID-19 vaccine (BNT162b2). Paired serum and breast milk samples were simultaneously taken from each participant at three timepoints after receiving the vaccine: 2 weeks after 1^st^ dose, 2 weeks after 2^nd^ dose and 4 weeks after 2^nd^ dose (Timepoints 1, 2 and 3, respectively). Levels of IgG antibodies against the spike protein (S1 subunit) were determined for each sample (Architect, Abbott®).

**Results:** we collected and analyzed 52 serum and 52 milk samples from the first 18 study participants. Median (interquartile range) IgG(S1) levels for serum – milk pairs at each timepoint were 410 (208-606) - 1.7 (0-2.9) AU/ml at Timepoint 1, 11505 (8933 - 21184) – 52.2 (34.1-113) at Timepoint 2 and 8311 (5578-17419) – 41.7 (24.8-75.3) at Timepoint 3. Pearson’s correlation coefficient between breast milk and serum IgG(S1) levels was 0.71. No major adverse reactions were observed in mothers or infants.

**Conclusions:** Breast milk from women vaccinated with mRNA-based Pfizer-BioNTech® vaccine contains specific anti-SARS-CoV-2 IgG(S1) antibodies, with levels increasing considerably after second dose. IgG(S1) levels in breast milk are positively correlated with corresponding serum levels.

## INTRODUCTION

The current COVID-19 pandemic caused by SARS-CoV-2 has caused great concern among breastfeeding women, both because of the possibility of viral transmission to infants during breastfeeding and, more recently, of potential harmful effects of vaccination in this specific population.

At the beginning of the vaccination campaign in our center, multiple healthcare professionals who were breastfeeding consulted our Infectious Disease Department, concerned about vaccination against COVID. Their main concern was whether breastfeeding was a contraindication to vaccination and, therefore, they felt they had to choose between being vaccinated or ending breastfeeding, since they were considered at high risk of infection. Although pregnant and lactating women have not been included in the clinical trials for the approval of these vaccines, the main official bodies and scientific associations consider mRNA vaccines as low risk for lactation and, therefore, they recommend its administration in cases where the risk of contracting the disease may be higher than the potential risks of vaccination^1-5^.

Recent studies have shown that milk produced by infected mothers is a source of anti-SARS-CoV-2 antibodies^6^, and two reports have demonstrated the existence of anti-SARS-CoV-2 antibodies in women vaccinated with Moderna® and Pfizer-BioNTech® mRNA vaccines^7,8^. If this exerts a protective effect for breastfed infants remains to be established.

In the present study, we characterize the levels of specific SARS-CoV-2 antibodies in the breast milk of mRNA-vaccinated women across time, as well as their correlation with serum antibody levels.

## METHODS

This prospective study included lactating women aged over 18 who were vaccinated against SARS-CoV2 with the Pfizer-BioNTech® COVID-19 vaccine (BNT162b2). The study received approval from our institution’s Ethics Committee (*CEIm Fundació Sant Joan de Déu*) with approval code PIC-28-21. All participants signed the corresponding informed consent.

Serum and breast milk samples were simultaneously taken from each participant at three timepoints: after receiving the first dose of the vaccine (timepoint 1), at two weeks-(timepoint 2) and four weeks after second dose (timepoint 3). At the same time, all participants underwent a nasopharyngeal smear for SARS-CoV-2 rapid antigen test determination (Ag-RDT). Levels of IgG antibodies against the spike protein (S1 subunit) and against the nucleocapsid (NC) of SARS-CoV-2 were determined for each sample (Architect, Abbott®). All statistical analyses were performed with R version 4.0.3 (2020-10-10) and figures were generated using the *ggplot2* R package.

## RESULTS

Here we show the results for the first 18 study participants. None of them had had confirmed SARS-CoV-2 infection prior to vaccination, nor during the study period (IgG-NC and Ag-RDT were all negative). Mean(SD) age and postpartum time were 37.8(3.1) years and 18.7(10.4) months, respectively. We collected and analyzed 52 serum and 52 milk samples from the 18 participants (two pair of samples could not be collected, one at timepoint 1 and another at timepoint 3). Samples from timepoint 1 were taken at a median (range) of 14 (8-21) days after the first dose, while samples of timepoints 2 and 3 were taken at 14 (10-18) days and 28 (26-32) days after the second vaccine dose, respectively.

Median (interquartile range) IgG(S1) levels for serum – milk pairs at each timepoint were 410 (208-606) - 1.7 (0-2.9) AU/ml for timepoint 1, 11505 (8933 - 21184) – 52.2 (34.1-113) for timepoint 2 and 8311 (5578-17419) – 41.7 (24.8-75.3) for timepoint 3 (Figure 1). Pearson’s correlation coefficient between breast milk and serum IgG(S1) levels was 0.71 (Figure 2).

**Figure 1.**
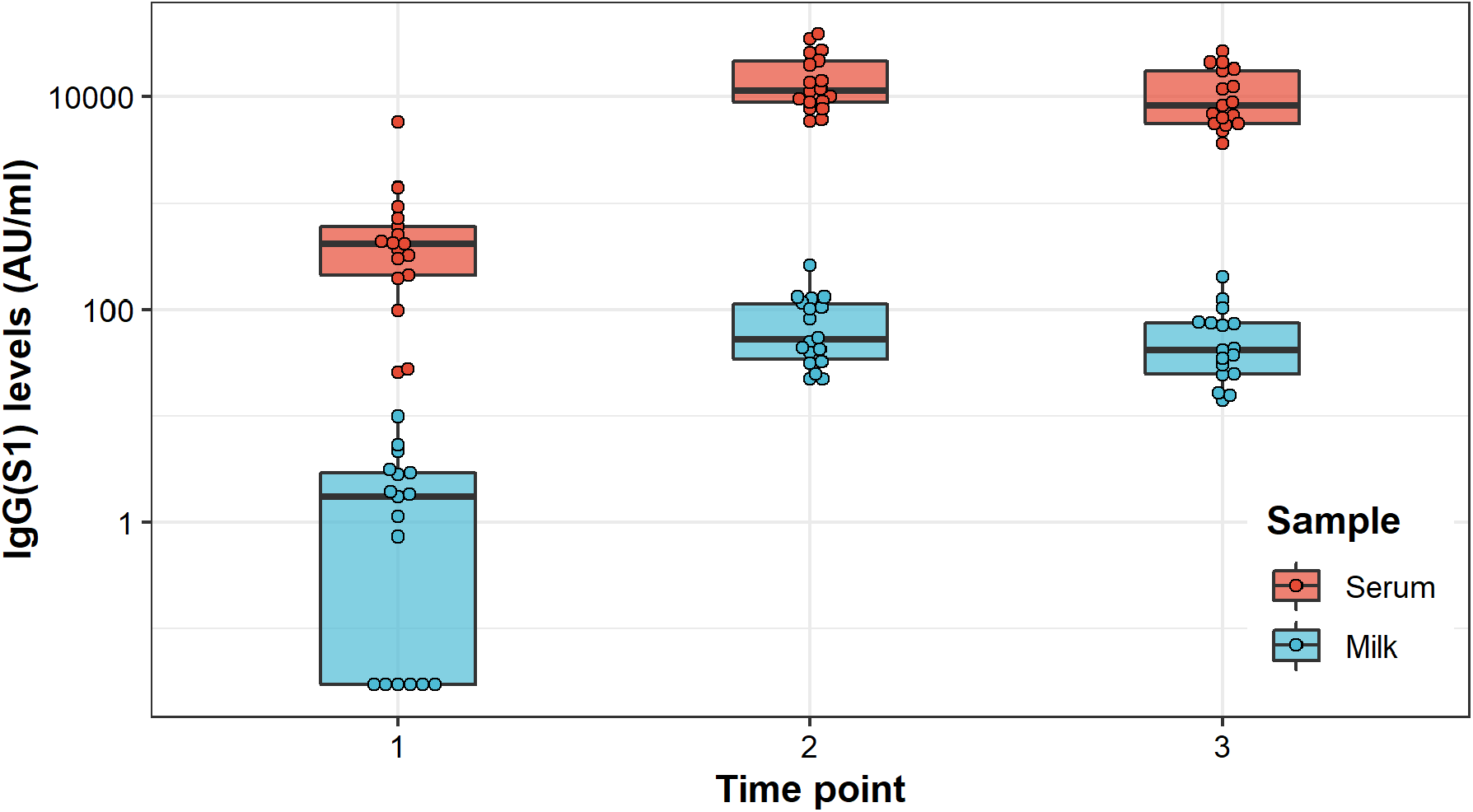
Evolution of IgG(S1) levels in Breast Milk and Serum of vaccinated participants across time. Box- and swarmplots showing superimposed serum (red) and breast milk (blue) IgG(S1) levels at the different timepoints. The scale on the Y-axis was transformed to log10 for visualization purposes and an offset of 0.03 was added to IgG1(S1) values to include 6 participants with undetectable antibody levels.

**Figure 2.**
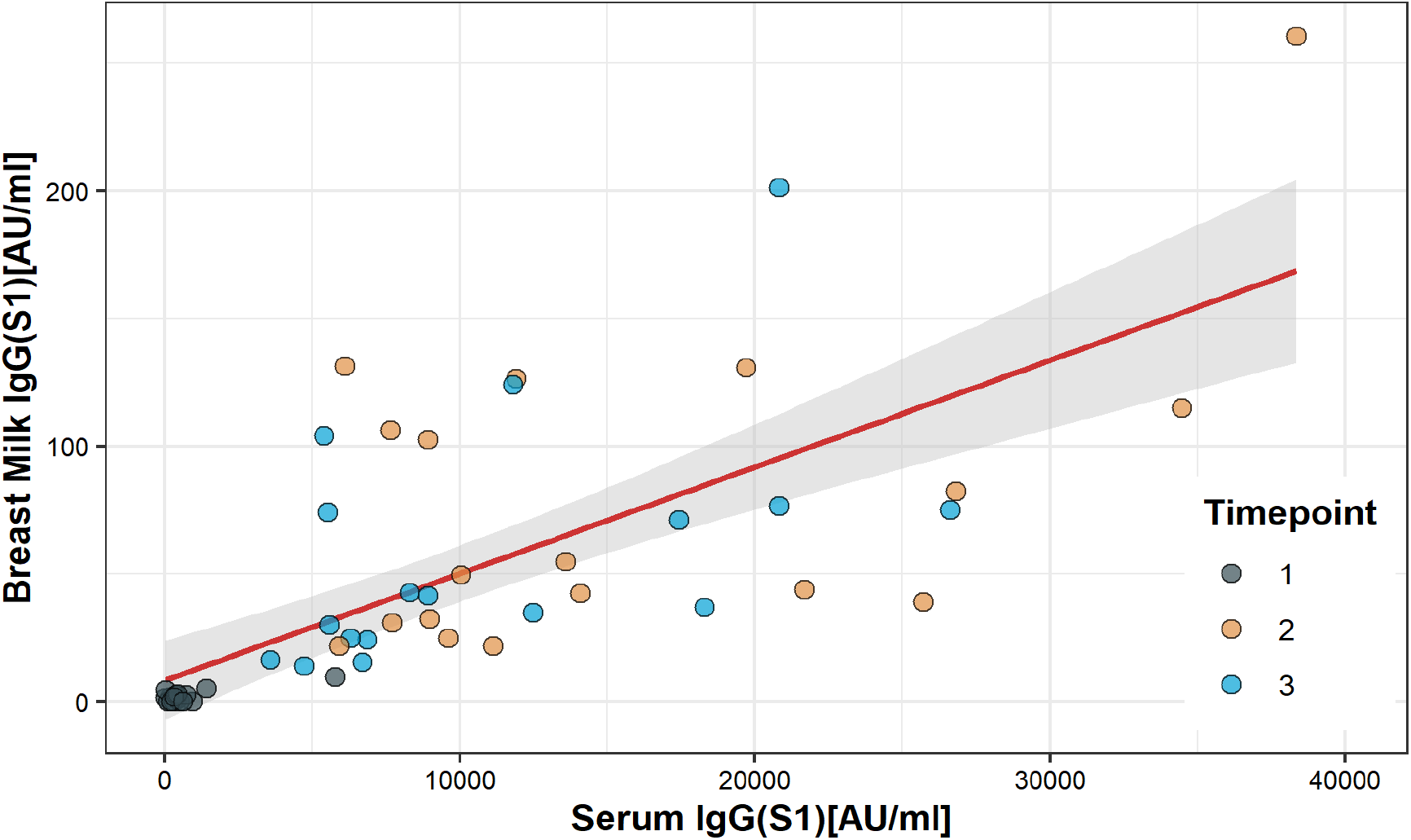
Correlation between IgG(S1) levels in Serum and Breast Milk of vaccinated participants. Scatterplot showing IgG(S1) levels for all samples, colored by timepoint. The regression line is shown in red, with 95% confidence intervals shaded in light gray.

All adverse reactions observed in mothers were minor. No adverse effects were observed in any of the lactating infants.

## DISCUSSION

Here we demonstrated that breast milk from women vaccinated with the novel mRNA-based Pfizer-BioNTech® vaccine contains specific anti-SARS-CoV-2 IgG(S1) antibodies. Furthermore, we showed that breast milk IgG(S1) levels dramatically increase after the second dose and that they are positively correlated with corresponding serum levels. This promising finding argues in favor of a potential protective effect of breastfeeding mothers’ vaccination in their infants. It remains to be determined if breast milk antibody levels decrease or plateau after vaccination, or whether these findings can be reproduced for other mRNA and non-mRNA-based vaccines. The kinetics of IgG and other specific immunoglobulins against SARS-CoV-2 such as IgA and IgM have been well studied after the disease^13^, mainly in serum but also in breast milk^14^, although their dynamics after vaccination remain unknown. Larger prospective studies examining these issues are needed to confirm the safety of SARS-CoV-2 vaccination in breastfeeding women and its impact on their infants’ health and SARS-CoV-2-specific immunity. Regarding the present study, accrual is still ongoing, and we plan to publish the results of more in-depth analysis in an expanded cohort of participants.

## CONCLUSIONS

Breast milk from women vaccinated with mRNA-based Pfizer-BioNTech® vaccine contains specific anti-SARS-CoV-2 IgG(S1) antibodies, with levels increasing considerably after second dose. IgG(S1) levels in breast milk are positively correlated with corresponding serum levels. Breastfeeding could have a protective effect against COVID-19 for breastfed infants with vaccinated mothers.

## Data Availability

Data will be available after study completion and upon request.

## Author Contributions

Concept and design: Esteve-Palau, Diaz-Brito.

Recruitment of volunteers and laboratory samples collection: Alvarez, Garcia-Aranda, Garcia-Terol, Esteve-Palau.

Laboratory samples analysis: Gonzalez-Cuevas, Guerrero.

Acquisition, analysis, or interpretation of data: All authors.

Drafting of the manuscript: Esteve-Palau, Casadevall, Diaz-Brito.

Critical revision of the manuscript for important intellectual content: All authors.

Statistical analysis: Esteve-Palau, Casadevall.

Supervision: Diaz-Brito.

All authors have seen and approved the manuscript.

## Acknowledgments

No compensation was received by any author for their role in this study, and all of them declare no conflicts of interest.

## Notes

### Competing Interest Statement

The authors have declared no competing interest.

### Author Declarations

The study received approval from our institution Ethics Committee (CEIm Fundacio Sant Joan de Deu) with approval code PIC-28-21. All participants signed the corresponding informed consent.

## REFERENCES

1. Paul E Sax. Covid-19 Vaccine Frequently Asked Questions. https://www.nejm.org/covid-vaccine/faq

2. http://www.e-lactancia.org/breastfeeding/covid-19-vaccine/product/

3. https://vacunasaep.org/profesionales/noticias/vacunacion-de-la-covid-en-el-embarazo-y-lactancia

4. https://www.ihan.es/comunicado-ihan-vacunacion-frente-a-covid-19-y-lactancia-materna/

5. https://blog.lactapp.es/compatibilidad-vacuna-covid-19-lactancia/

6. Pace RM, Williams JE, Järvinen KM, et al. Characterization of SARS-CoV-2 RNA, Antibodies, and Neutralizing Capacity in Milk Produced by Women with COVID-19. mBio. 2021;12(1):e03192–20.

7. Jill K Baird, Shawn M Jensen, Walter J Urba et al. SARS-CoV-2 antibodies detected in human breast milk post-vaccination. Preprint medRxiv 2021.02.23.21252328. https://doi.org/10.1101/2021.02.23.21252328

8. Michal Rosenberg Friedman, Aya Kigel, Yael Bahar et al. BNT162b2 COVID-19 mRNA vaccine elicits a rapid and synchronized antibody response in blood and milk of breastfeeding women. https://doi.org/10.1101/2021.03.06.21252603

9. Stuebe A. The risks of not breastfeeding for mothers and infants. Rev Obstet Gynecol 2009;2(4):222–31.

10. Edmond KM, Zandoh C, Quigley MA, Amenga-Etego S, Owusu-Agyei S, Kirkwood BR. Delayed breastfeeding initiation increases risk of neonatal mortality. Pediatrics 2006;117(3):e380–6.

11. Smith ER, Locks LM, Manji KP, et al. Delayed breastfeeding initiation is associated with infant morbidity. J Pediatr 2017;191:57–62.e2.

12. NEOVITA Study Group. Timing of initiation, patterns of breastfeeding, and infant survival: Prospective analysis of pooled data from three randomised trials. Lancet Glob Health 2016;4(4):e266–75

13. Deeks JJ, Dinnes J, Takwoingi Y, et al. Antibody tests for identification of current and past infection with SARS-CoV-2. Cochrane Database Syst Rev. 2020;6:CD013652.

14. Demers-Mathieu V, Do DM, Mathijssen GB, et al. Difference in levels of SARS-CoV-2 S1 and S2 subunits- and nucleocapsid protein-reactive SIgM/IgM, IgG and SIgA/IgA antibodies in human milk. J Perinatol. 2020 Sep 1;1–10. https://doi.org/10.1038/s41372-020-00805-w.

